# The effect of opening up the US on COVID-19 spread

**DOI:** 10.1101/2020.07.03.20145649

**Authors:** Patrick Bryant, Arne Elofsson

## Abstract

In response to the pandemic development of the novel coronavirus (SARS-CoV-2), governments worldwide have implemented strategies of suppression by non-pharmaceutical interventions (NPIs). Such NPIs include social distancing, school closures, limiting international travel and complete lockdown. Worldwide the NPIs enforced to limit the spread of COVID-19 are now being lifted. Understanding how the risk increases when NPIs are lifted is important for decision making. Treating NPIs equally across countries and regions limits the possibility for modelling differences in epidemic response, as the response to the NPIs influences can vary between regions and this can affect the epidemic outcome, so do the strength and speed of lifting these. Our solution to this is to measure mobility changes from mobile phone data and their impacts on the basic reproductive number. We model the epidemic in all US states to compare the difference in outcome if NPIs are lifted or retained. We show that keeping NPIs just a few weeks longer has a substantial impact on the epidemic outcome.

## Introduction

In response to the pandemic development of the novel coronavirus (SARS-CoV-2), governments worldwide implemented strategies of suppression by non-pharmaceutical interventions (NPIs). Such NPIs include social distancing, school closures, limiting international travel and the strongest being complete lockdown^1,2^. The goal of the NPIs is to reduce the spread of the infection by diminishing the basic reproductive number R, which represents the average number of infections each infected individual will create. Spread is expected to continue if R>1 and end if R<1^3^.

The NPIs implemented to limit the spread of COVID-19 are now being lifted worldwide ^4^. The effect of the opening up will influence each country and region differently depending on parameters such as (1) the strength of the initial NPIs (2) the number of daily infections at the opening (3) the cumulative number of infections at the opening (4) the speed and (5) the strength of the opening. All of these parameters will work together to influence the final outcome.

Understanding what parameters to track and the difference in outcome as a result of when, how much and how fast NPIs are lifted is important for decision making. Treating the effect of NPIs equally across countries and regions^5^ limits the possibility for modelling differences in response and thus the outcome. Instead, the effect of NPIs and their lifting can be traced using region-specific mobility data^6^ from mobile phones.

At the beginning of May 2020, opening began across US states, at a countrywide level of above 1 million known infections and over 60 000 deaths ^4^. The opening, as the NPI implementation in mid March^2^, was accompanied by rapid mobility changes across all the indicators of mobility except for the parks, which displays more dubious cyclic behaviour (Figure 1).

**Figure 1.**
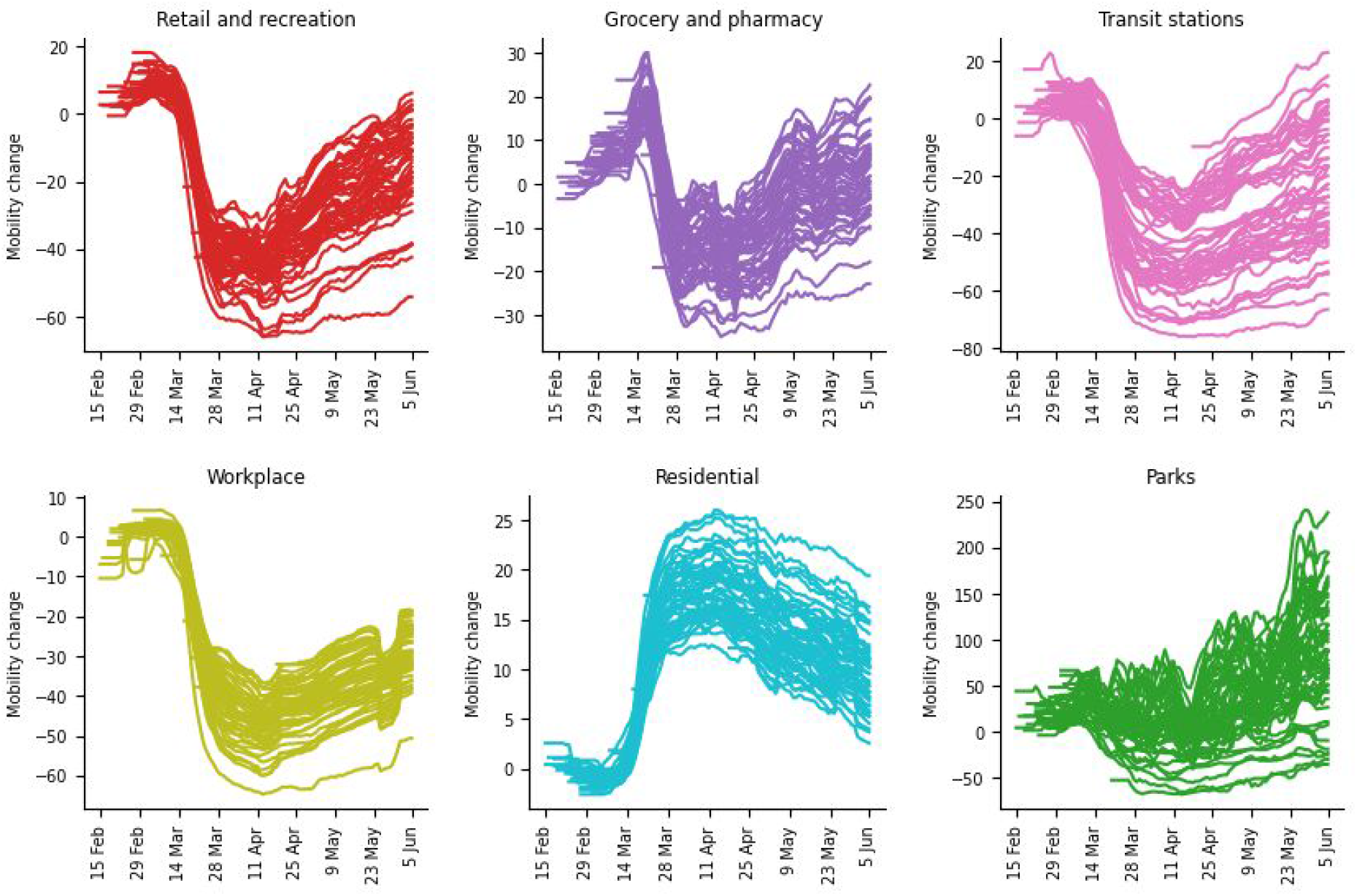
Mobility changes in all 50 US states in the six sectors, retail and recreation, grocery and pharmacy, transit stations, workplace, residential and parks, reported by google mobility analysis. The mobility data is smoothed using a one-week sliding average. The mobility change is the value change compared to a baseline estimated median value, for each day, during the period 2020-01-03 to 2020-02-06^6^. Mobility in the park sector displays no clear lockdown response and appears to have a cyclic tendency, as of following the weather and/or public holidays, therefore this data was not used in our model.

By using data about mortality in Covid19 for each US state from Johns Hopkins University^7^, we model the impact of this opening in a Bayesian posterior distribution as a function of the number of estimated infections. Our model estimates the impact on the basic reproductive number caused by mobility changes. To ensure the relationship between mobility changes and R is true and not a model artefact, we estimate R independently from cases using the package EpiEstim^8^ for comparison. The outcomes from the mobility changes are compared with the hypothetical scenario of a continued lockdown, simulated by maintaining the mobility levels observed at the extreme points following NPI implementations.

## The impact of introducing non-pharmaceutical interventions in the USA

There is a clear relationship between the mobility changes and independent estimates of basic reproductive numbers (R) before and after NPI introduction (Figure 2). This relationship is nonlinear, having a sharp drop in R at certain mobility thresholds. The correlations are similar for all categories (≈ 0.7). The states with strong NPI responses (e.g. New York, Louisiana and Michigan) show an almost complete reduction in spread with R values under 1 (Figure 3), while in states where the lockdown was not very strong (e.g. North Carolina), a flatter curve with continued spread (R close to, but still above 1) is observed.

**Figure 2.**
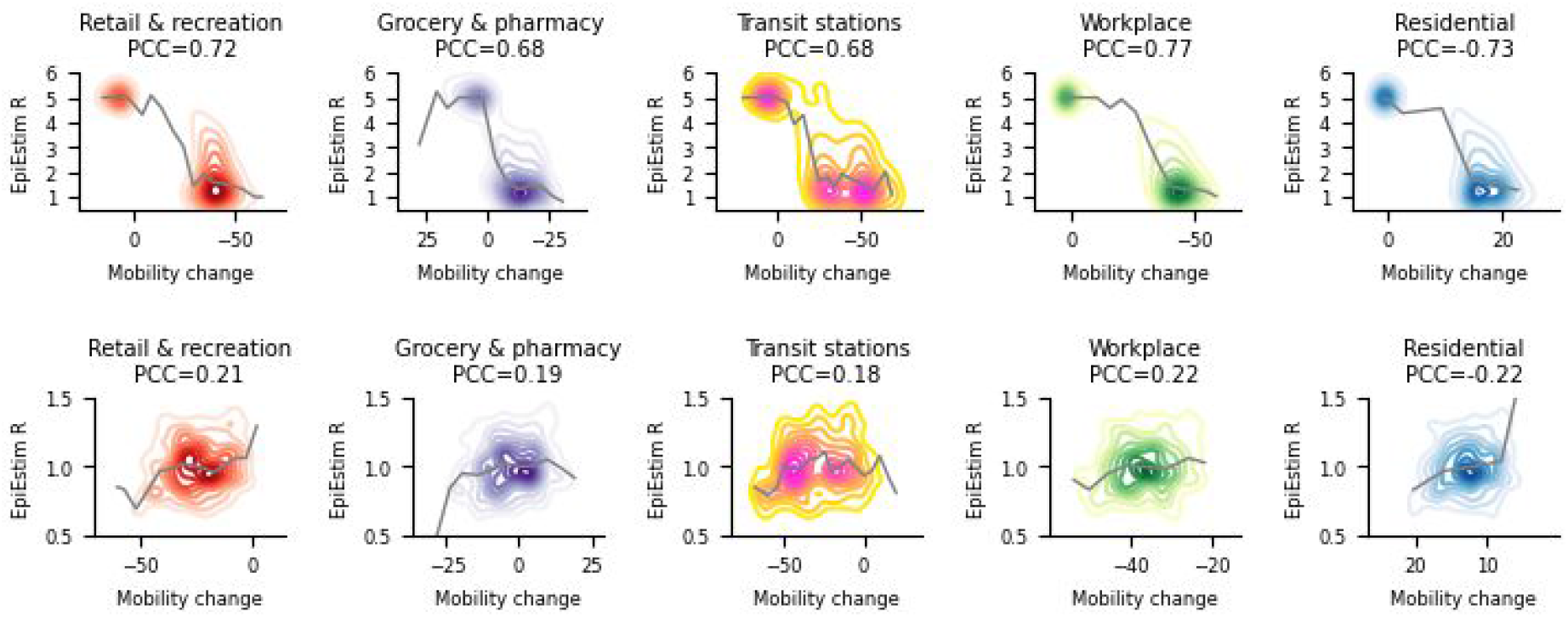
Relationship between the mean R from independent case estimates, using the EpiEstim package^8^, and mobility changes in all states as kernel density plots. The thin grey lines represent the median relationship in intervals of 5 % mobility change. The top row shows the values from the epidemic start to after NPI implementation (April 25). The bottom row shows the minimum values from after NPI implementation (April 25) to after the NPI lifting (June 5). The Pearson correlation coefficients (PCC) are marked for each category.

**Figure 3.**
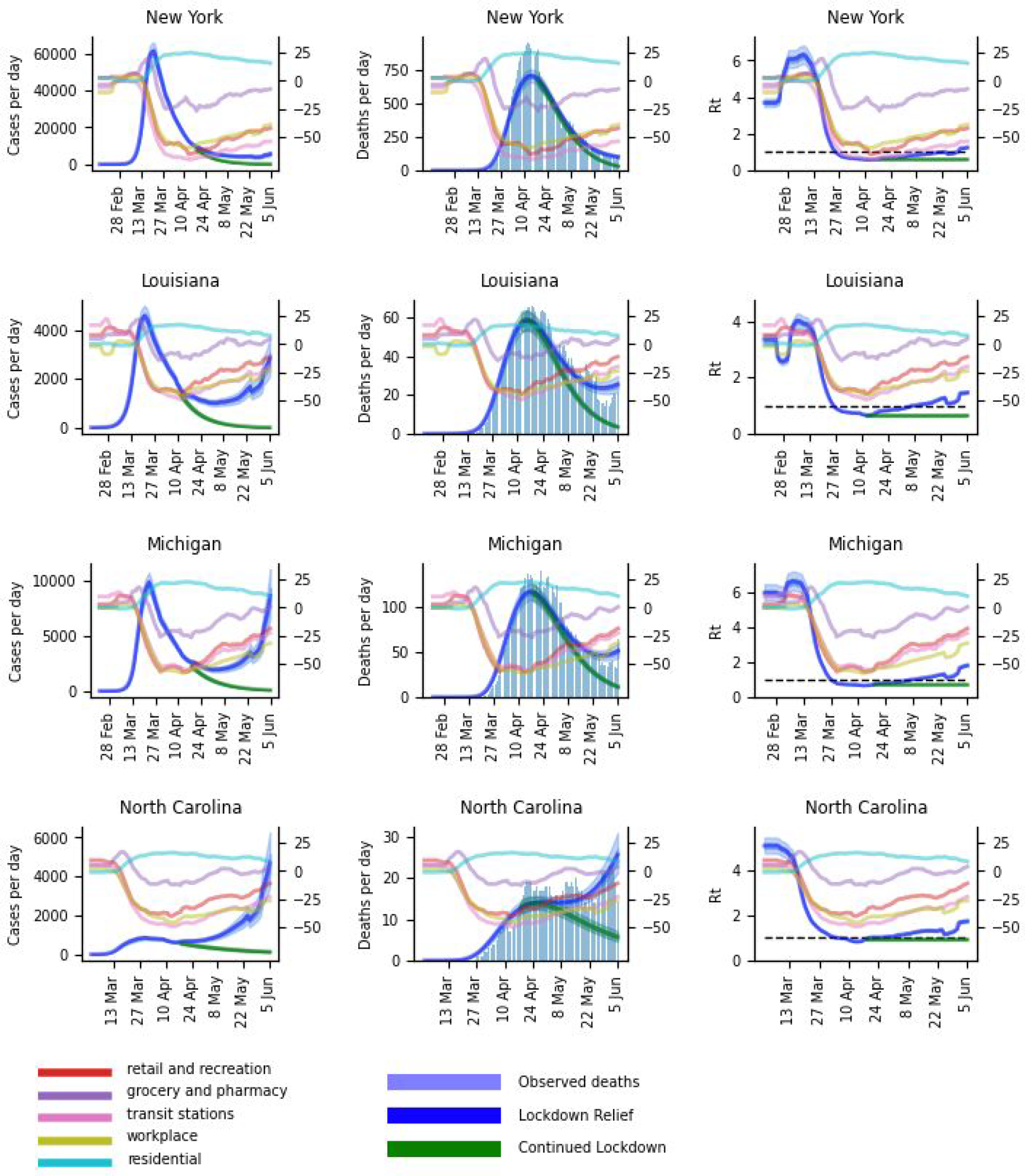
The impact of opening up each state displayed as daily cases, deaths and basic reproductive number (Rt). The blue curve represents the actual modelled outcome, the green the outcome upon continued lockdown (from the intersection of the two curves) and the light blue histogram the actual reported deaths. The 50 and 95 % CIs are marked in lighter shades of blue and green respectively. The mobility changes from the 5 sectors are marked as thin lines. The graph for Rt includes a dashed line marking the value 1 of halted epidemic growth. All data is smoothed using a one-week sliding average.

## The impact of lifting non-pharmaceutical interventions in the USA

The relationship between the mobility changes and independent estimates of R before and after NPI lifting (Figure 2) show less clear correlations than that of NPI introduction. Similar relationships are displayed though, with all but the residential sector having a positive correlation with R. The correlations per sector are low (≈ 0.2). All estimated R values here are very close to 1 and the variation displayed is therefore prone to uncertainty in these estimates.

The states which maintained their strong NPI responses (e.g. New York) show narrow peaks in daily deaths and cases (Figure 3). There, the NPI relief has had little impact on the outcome, since the mobility has been kept at low levels, resulting in an R-value of close to 1. Other states (e.g. Louisiana and Michigan) have had moderate NPI reliefs, resulting in flatter curves, with R-values slightly above 1. In states where the lockdown was not very strong, but the opening rather stronger and faster (e.g. North Carolina), an upward trend in daily cases (R closer to 2) and deaths is observed.

In states where the mobility changes were not maintained, a resurgence in spread is observed (Table 1). By prolonging the NPIs only a few weeks more, a completely different starting point can be obtained for NPI relief. This can be observed especially in Louisiana, Michigan and North Carolina (green curve, Figure 3). If the NPIs would have been retained the fractional deaths of the previous peak would be 4.6 % vs 14.2 % with the lifting in New York, but 6.9 vs 43.1, 10.3 vs 43.6 and 42.9 vs 185.7 % without and with NPI lifting for Louisiana, Michigan and North Carolina respectively.

**Table 1.** Deaths modelled with NPI lifting, NPI continuation and their percentage of the previous death peak at June 5 with 95 % confidence intervals.

| State | Modeled deaths with NPI lifting | Percent of previous death peak with NPI lifting | Modeled deaths with continued NPIs | Percent of previous death peak with continued NPIs |
| --- | --- | --- | --- | --- |
| New York | 100.0 [87.0,115.0] | 14.2 % [12.4,16.4] | 32.0 [28.0,37.0] | 4.6 % [4.0,5.3] |
| Louisiana | 25.0 [21.0,30.0] | 43.1 % [36.2,51.7] | 4.0 [3.0,4.0] | 6.9 % [5.2,6.9] |
| Michigan | 51.0 [44.0,60.0] | 43.6 % [37.6,51.3] | 12.0 [10.0,13.0] | 10.3 % [8.5,11.1] |
| North Carolina | 26.0 [21.0,31.0] | 185.7 % [150.0,221.4] | 6.0 [5.0,7.0] | 42.9 % [35.7,50.0] |

All states display similar relationships (Figure S1, Table S1), with spread following mobility increase and both strength and timing of NPI introduction and lifting greatly affecting the outcome. Most states resemble Louisiana and Michigan, having curves that increase in cases but flatten out in deaths upon NPI lifting. Some tend more to that of North Carolina, while only Washington resembles that of New York.

## Discussion

As non-pharmaceutical interventions (NPIs) are lifted across the USA we model the impact of the timing and strength of the lifting on the epidemic outcome through changes in mobility patterns. The mobility data across all states are very similar, still there are differences which have a substantial impact on the outcome. Treating NPIs equally across countries and regions^5^ limits the possibility for modelling differences in epidemic response, as the NPI implementation strength influences the epidemic outcome, so do the strength and speed of lifting these.

Since the number of known cases is thought to be underestimated and the undocumented cases account for a large portion of spread ^9^, the number of reported daily deaths are modeled in a Bayesian posterior distribution as a function of the number of estimated infections. The estimations are sensitive to reporting delays and spurious peaks, such as observed on May 8 (Figure S1). Uneven reporting, due to e.g. policy change, is, therefore, a large source of error. Smoothing the data by a one-week running average reduces some of the uncertainty, but makes it harder to capture trends depending on future data. This can be observed when comparing Figures S1 and S2, which display smoothed and actual reported daily deaths respectively.

The independent R estimates from daily cases and their relationship with mobility changes give strong support for the usefulness of tracking mobility. The observations from NPI lifting are less clear than of NPI introduction, which is most likely due to the small changes in mobility and thus small changes in R. Small changes make the estimates less certain, as an error in estimating R of only 0.1 can mean the difference between exponential spread (R=1.1) and epidemic end (R=0.9).

All states are estimated simultaneously in a hierarchical framework, assuming the mobility changes have similar impacts across all states. Even though mobility changes can be useful proxies for tracking epidemic spread, other factors such as differences in population interconnectedness, infection fatality ratios^10^, social distancing, and use of personal protective equipment will affect the outcome. Differences in reporting and the amount of available data for each state will also contribute. These unknowns are not considered in the model, making some states display better model fits than others (Figure S1).

## Conclusions

The difference in outcome between NPI lifting and retention suggests large benefits with prolonging NPIs only a few weeks more and emphasizes the importance of not opening up too early or too fast. That such small costs can result in such large benefits is important to take into account for policymakers and those tracking the epidemic development.

## Methods

### Epidemic Model

We model the impact on the basic reproductive number (R) as a function of changes in mobility data for each state^11^, estimated simultaneously for all states in a hierarchical Bayesian framework using Markov-Chain Monte-Carlo (MCMC)^12,13^ simulations:

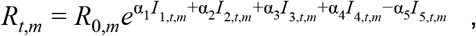

where I_1-5,t,m_ is the one-week sliding average in relative mobility in retail and recreation, grocery and pharmacy, transit stations, workplace and residential sectors respectively at day t in state m. The prior for R_0_ ^9,14^ is set to:

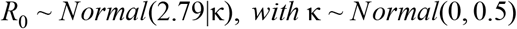

The relative mobility is the value change compared to a baseline estimated median value, for each day, during the period 2020-01-03 to 2020-02-06^6^.

The number of daily deaths (D_t,m_) in each state is modeled in the Bayesian posterior distribution as a function of the cumulative number of cases from the previous days, weighted by a serial interval distribution, times R at day t in a negative binomial distribution^15^.

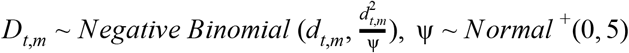

The expected number of deaths, d_t,m_, at day t in state m is given by:

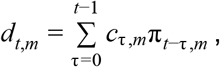

where π_*m*_ is the infection to death distribution in state m given by a combination of the infection to onset distribution (Gamma(5.1,0.86)) and onset to death distribution (Gamma(18.8,0.45)) (combined with mean 23.9 days and standard deviation 0.45 days) times the infection fatality rate (ifr), set to be 1 % for all states^16,17^ :

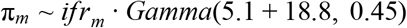

π_m_ is discretized in steps of 1 day: 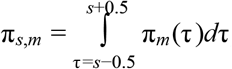 *for s* = 2, 3, … *and* 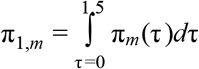

The number of cases acquired art day τ in country m, *c*_*τ,m*_ is modelled with a discrete renewal process ^18,19^

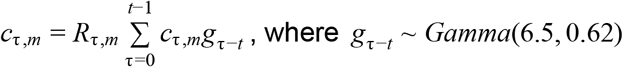

The serial interval distribution used to model the number of cases.

g_s_ is discretized in steps of 1 day: 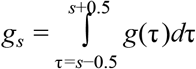 *for s* = 2, 3, … *and* 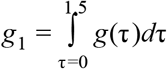

We start the simulation for each state 30 days before the day after 10 observed cumulative deaths. For stability, we initialize our model with 6 days^9^ of cases drawn from an Exponential(0.03) distribution, which are inferred in the Bayesian posterior distribution (D_t,m_).

The death data was reported from John Hopkins University^7^. We ran the sampling with eight chains, using 4000 iterations (2000 warm-up), as in earlier work^5,13^, ensuring no divergent transitions were observed (Figure S3). The parameter specifics of the simulation are available in the code.

### Estimating the outcome of continued non-pharmaceutical interventions

To estimate the outcome of continued NPIs, we select the most extreme mobility values observed after NPI implementation in each state. The values for the date when these extremes had been reached across all five mobility categories were used to model the alternative outcome (Figure S4).

By selecting the estimated R values from the initial simulation using the observed mobility changes, we calculate the alternative outcome using the same discrete renewal process described above.

### Estimating the basic reproductive number with EpiEstim

To ensure there truly are relationships between spread and mobility changes, we estimate R independently using case data from John Hopkins University ^20^ and the R package EpiEstim^8^. The R estimates are uncertain in the beginning of the epidemic when cases are few, why they are smoothed using one-week averages and only values under 6 are included (Figure S5). The serial interval used for the estimations is variable accordingly: estimate_R(country_cases, method=“uncertain_si”, config = make_config(list(mean_si = 7.5, std_mean_si = 2,min_mean_si = 1, max_mean_si = 8.4, std_si = 3.4, std_std_si = 1, min_std_si = 0.5, max_std_si = 4, n1 = 1000, n2 = 1000))), allowing more possible scenarios to be explored.

### Code

https://github.com/patrickbryant1/COVID19.github.io/tree/master/simulations/mobility/dev/US

## Data Availability

All code and data are freely available.

https://github.com/patrickbryant1/COVID19.github.io/tree/master/simulations/mobility/dev/US

## Author Contributions

PB and AE jointly designed the study. PB performed the experiments and wrote the initial draft, which was further edited, revised and approved by both authors.

